# Differences in lower extremity joint coordination during two landing phases of a drop jump task

**DOI:** 10.1101/2023.07.22.23292930

**Authors:** Jia-Wei Wang, Ye Liu

## Abstract

**Objective:** The aim of the present study was to compare the differences in joint coordination patterns and variability of the lower extremity between the first and second landing phases during drop jump task.

**Design:** Cross-sectional study.

**Methods:** Modified vector coding technique and circular statistics was used to determine the coordination pattern and variability of the following joint couples during the first and second landings: hip frontal-knee frontal (HfKf), hip sagittal-knee frontal (HsKf), hip sagittal-knee sagittal (HsKs), knee frontal-ankle frontal (KfAf), knee sagittal-ankle frontal (KsAf), and knee sagittal-ankle sagittal (KsAs).

**Results:** The second landing phase exhibited a reduction in the in-phase coordination proportion of HsKs, KfAf, and KsAs, while demonstrating an increase in the proportion of proximal knee joint coordination for KfAf and KsAs (P<0.05). The second landing phase demonstrated increased coordination variability for HsKs, KfAf, KsAf.

**Conclusion:** The execution of the drop jump leads to changes in joint coordination patterns during the second landing phase, resulting in increased variability compared to the first landing phase, thereby elevating the risk of knee and ankle injuries.

## 1 Introduction

The drop jump (DJ) is commonly utilized as a screening exercise for lower extremity injury risk factors^1, 2^. It involves a sequence of actions, including descending from a raised platform, the first landing, subsequent maximal vertical jump, and the second landing. During each landing, the stress exerted on various joints of the lower extremity can lead to various non-contact sports injuries, such as anterior cruciate ligament (ACL) injuries^3^. These types of injuries are particularly common in sports involving repetitive jumping movements. Therefore, the DJ serves as a valuable tool for simulating common injury mechanisms and offers the advantage of high levels of reliability^4^.

Currently, there is a significant emphasis on the biomechanical analysis of the first landing in DJ, while research investigating the second landing following the maximal vertical jump is limited. The initial contact with the ground in DJ, known as the “first landing,” is controlled through the provision of visual and auditory instructions to participants, enabling them to focus more on their own movements. Conversely, participants are not given explicit instructions before the subsequent contact with the ground, referred to as the “second landing,” and tend to prioritize external or non-specific goals. ACL injuries frequently occur during the second landing phase after the maximal vertical jump in various sports^5, 6^. Therefore, compared to the first landing in DJ, the second landing can more accurately simulate the mechanisms of injury risk. Previous studies have indicated that different motion patterns involve distinct neuromuscular strategies and carry varying risks of sports-related injuries^7, 8^. Researchers have identified fundamental biomechanical discrepancies between the first and second landing phases in DJ. The second landing is characterized by a higher center of mass position^9^, increased vertical ground reaction force (GRF)^2^, greater side-to-side asymmetry, and an increased knee abduction angle^3^, in contrast to the first landing. Consequently, the second landing involves less favorable motion patterns and neuromuscular strategies, potentially increasing the risk of sports-related injuries upon landing. However, these studies have primarily focused on the biomechanics of single joints, without considering the relative motion between two joints. During weight-bearing activities, the human body exhibits complex interactions between joints^10^. Assessing abnormal motion patterns solely based on single-joint motion has inherent limitations. Therefore, it is crucial to analyze the coordination patterns between joints.

Joint coordination and its variability explain how the neuromuscular system integrates multiple limbs, joints, and senses to produce smooth, goal-oriented movements while maintaining flexibility to adapt to changes in the environment or task requirements^11^. According to Bernstein, due to the redundant number of degrees of freedom in human motion, no movement could be completely replicated^12^. The variability resulting from redundancy is often regarded as “noise” by traditional biomechanical analysis methods, which is useless or detrimental information^13^. With the development of dynamic systems theory, variability is recognized as a facilitator of individual motor learning. For example, novices typically exhibit lower variability during the execution of movements, while high-level athletes can employ more variability to accomplish the task^14^. Although a certain amount of variability during movement can facilitate adaptation to task demands or environmental changes, increased variability in highly controlled environments, such as on flat ground or during undisturbed activities, may indicates a motor control system instability than superior flexibility of the neuromuscular system^11^. This can place individuals in abnormal biomechanical patterns and high-risk joint loading. Additionally, based on the variability-overuse injury hypothesis, reduced coordination variability may lead to repeated loading of specific tissues, resulting in overuse injuries^15^. The extensive discussions surrounding variability make it challenging to define its nature precisely. Therefore, it is meaningful to explore joint coordination variability that may carry potential injury risks. The Modified vector coding technique, based on dynamic systems theory, is a widely used method that quantitatively assesses the coordinated motion and variability between two joints. It can provide sensitive information related to injury mechanisms^16^. For example, Nordin et al. calculated the Lower extremity variability during DJ at different heights and found that variability decreased with increasing height^17^. Mah et al. demonstrated that sleep restriction increased Lower extremity variability in athletes performing DJ^18^. Considering previous research, discussions on joint coordination variability during the landing phase of DJ have primarily focused on the first landing. Therefore, it is necessary to adopt a dynamic systems approach to comprehensively analyze the coordination and variability between lower extremity joints during both pre– and post-landing phases of DJ task.

The objective of this study is to compare the coordination patterns and variability of the lower extremity joints during the first and second landing phases of a DJ task. Specifically, our aim is to investigate potential differences in the four coordination patterns and variability between these two landing phases. Previous research has indicated that the second landing demonstrates more higher rigor task compared to the first landing. Therefore, we hypothesized that there are disparities in coordination patterns between the first and second landing phases, with a greater proportion of anti-phase coordination observed during the second landing. Moreover, considering the connection between landing-related injury risk and coordination variability, we propose that the second landing will exhibit higher variability and an increase in side-to-side asymmetry compared to the first landing.

## 2 Methods

This study utilized a cross-sectional research design to examine differences in lower extremity joint coordination between the first and second landing phases of DJ task performed by adult male athletes. Each participant completed five DJs from a height of 0.4m. The participants were experienced adults who participated in resistance training, including plyometric exercises, at least three times per week. The choice of a 40 cm height for the DJ was informed by previous research, which frequently examined lower extremity biomechanical differences at this specific height^2^.

### Participants

This study recruited 18 resistance-trained adult participants (age: 22.8±1.8 years; height: 1.79±0.62 m; mass: 75.0±6.5 kg) who volunteered to take part in the research. The study received ethical approval from the Academic Division of Sports and Health of the Beijing Sport University (approval number: 2021171H; approval date: November 19, 2021). The inclusion criteria were as follows: (1) males between the ages of 18 and 25 years; (2) no history of lower extremity injuries, neurological impairments, vestibular system disorders, visual impairments, or any other relevant diseases within the past 3 months; (3) no intense physical activity or muscle fatigue within 72 hours prior to testing. The participants reported engaging in regular resistance training for a minimum of 2 years (defined as a minimum of 3 sessions per week), and had an average 1-repetition maximum parallel back squat of 143 kg (range: 120-185 kg). Before the experiment, all participants received comprehensive information regarding the experimental procedures and potential risks. They provided their informed consent prior to participating in the study.

### Experimental protocol

An eight-camera motion capture system (200Hz, Vicon Motion Analysis, Inc., Oxford, UK) was utilized to record kinematic trajectories. Synchronized data at 1000Hz were concurrently obtained using two force plates (9287C, Kistler Instruments AG Corp., Winterthur, Switzerland) with dimensions of 1.75×0.5 meters. To ensure consistency, a single examiner affixed 37 retroreflective markers, each with a diameter of 10mm, to predetermined anatomical landmarks. Reflective markers were placed the lower back between the S5 and T1 vertebrae, and bilaterally on the acromion processes, lateral epicondyle of the elbow, distal end of the forearm midway between the styloid processes of the radius and ulna, anterior superior and posterior inferior iliac spines, greater trochanter, medial and lateral femoral condyles, tibial tubercle, medial and lateral malleoli, anterior midthigh, lateral and anterior distal aspects of the shank, heel, dorsal surface of the midfoot, and central forefoot between the second and third metatarsals. With the exception of the markers attached to the shoes, all other markers were directly affixed to the skin. Static calibration data were subsequently collected after applying the markers. In order to minimize the influence of footwear-related factors, standardized socks and shoes of the same brand and material were worn by all participants.

The experiments were conducted in an indoor laboratory. Participants were instructed to complete a standardized 10-minute warm-up. Additionally, participants were given sufficient opportunity to practice DJ and become familiarize with the test procedures. Each participant, under the supervision and guidance of the examiner, performed five DJs from a height of 0.4m, with 15-20 seconds of rest between each attempt. The entire testing session lasted approximately 15 minutes. Before each jump, participants were verbally instructed to stand naturally on the box, both hands had to be held akimbo (hands at the ilium), and their feet were positioned approximately 0.3m apart with toes pointing forward. After receiving the “start” command from the examiner, participants lifted their dominant leg (the preferred limb used to kick a ball) naturally leaned forward, and executed a drop straight down using both feet from the 0.4m-high box, followed by a maximal vertical jump upon contact with the force plates. No specific instructions were provided for the execution of the second landing. They were required to land with both feet contained in separated force plates. If participants failed to land with both feet contained in separated force plates on the first and second landing then the trial was excluded from analysis.

### Data analysis

All data were imported into Visual3D Biomechanics Software suit (v6.0, C-Motion, Germantown, MD, USA) and a low-pass fourth-order Butterworth filter with a cut-off frequency of 12Hz was applied to process the marker trajectories^19^. An X-Y-Z (flexion-adduction-rotation) Cardan rotational sequence and the right-hand rule were used for 3D angular kinematics computations and polarizations, respectively. A 6-degrees-of-freedom skeletal model was utilized to analyze the filtered trajectories, allowing us to determine the position and orientation of each segment at each time sample. Additionally, the model was scaled based on the participants height and weight of each participant. Each landing phases was identified with kinematic and kinetic data. The first point was defined as the initial contact point with the force plate (vertical ground reaction force > 10N), and the end point was defined as the lowest point of the center of mass (CoM)^9^. The landing phase was time-normalized to 101 data points, representing 0% at the initial contact point and 100% at the lowest point of CoM.

A modified vector coding technique was employed to calculate the coupling angle (γ) and coupling angle variability (CAV). The couplings of interest included the following: hip frontal-knee frontal (HfKf), hip sagittal-knee frontal (HsKf), hip sagittal-knee sagittal (HsKs), knee frontal-ankle frontal (KfAf), knee sagittal-ankle frontal (KsAf), and knee sagittal-ankle sagittal (KsAs). Modified vector coding technique was performed using a custom-written MatLab code (MatLab 2022b, MathWarks, Inc., Natick, MA). which quantified the coordination date based on the following formula^20^:

The coupling angle (γ) at each instant (i) was calculated on the basis of proximal joint angle θ*_D_* and distal joint angle θ*_p_*, where *i* represents the trial number:

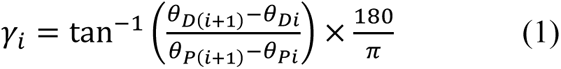

The mean coupling angle (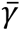) over three trials in each participant was calculated using circular statistics of Equations (2-4):

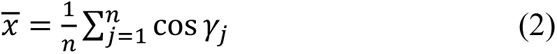

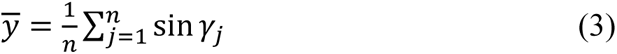

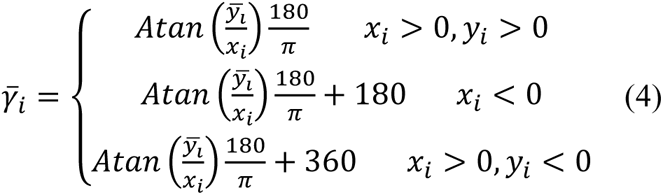

CAV was calculated according to (5):

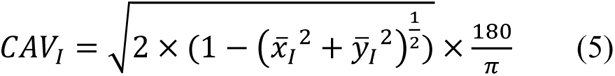

The mean coupling angle(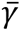) were categorized into one of four patterns within the range of 0-360°^21, 22^: (1)In-phase (112.5°≤θ<157.5°, 292.5°≤θ<337.5°): Two adjacent joints rotate in the same direction with similar amplitudes; (2)Anti –phase (112.5°≤θ<157.5°, 292.5°≤θ<337.5°): Two adjacent joints rotate in opposite directions; (3)Proximal phase (0°≤θ<22.5°, 157.5°≤θ<202.5°, 337.5°≤θ<360°): Proximal joint rotates dominantly compared to the distal joint; (4)Distal phase (67.5°≤ θ<112.5°, 247.5°≤θ<292.5°): Distal joint rotates dominantly compared to the proximal joint; The CAV was interpreted as the variation in participants’ landing strategies for each of the examined couplings. The average CAV values for each joint coupling during the landing phase of stance were submitted for statistical analysis^19^.

### Statistical analysis

Statistical analyses were performed with SPSS Statistics version 26.0(SPSS Inc., Chicago, USA). Normality was assessed using Shapiro–Wilk’s test and Levene’s test for equality of variances. Statistical differences in the distribution frequencies of coupling angles and CAV between the 2 lower extremity across the 2 landing phases were determined using a 2-way analysis of variance (ANOVA) (side-to-side [2-levels] × landing [2-levels]) with repeated measures on one factor(landing). Pairwise t-tests were used to calculate the 95% confidence limits (95% CL), and determine the p–values for differences within the ANOVA model. For all analyses, the significance level (α) was set at 0.05. Cohen’s d was computed to measure the effect size (ES) of differences, Cohen’s d and interpreted as follows: small>0.2; medium>0.5; large>0.8 ^23^.

## 3 Results

The proportions of lower extremity joint coordination patterns during different landing phases are depicted in Figure 1 and Figure 2. For HfKf couples, coordination patterns were concentrated in distal phase (knee rotates dominantly compared to the hip) on two landing phases. Notably, significant landing × side-to-side interactions were found for the in-phase(P=0.042). Specifically, the first landing demonstrated a significantly less proportion of in-phase on the nondominant limb compare with the second lan ding (P=0.035, ES=-0.73), whereas on the dominant limb, the proportion of in-phase during the first landing was significantly higher than nondominant (P=0.041, ES=0.72). Furthermore, a significant main effect for the factor of limb dominance on distal phas e (p<0.05), with the proportion of distal phase on the dominant limb during the first landing being significantly lower than nondominant (P=0.023, ES= –0.81).

**Figure 1.**
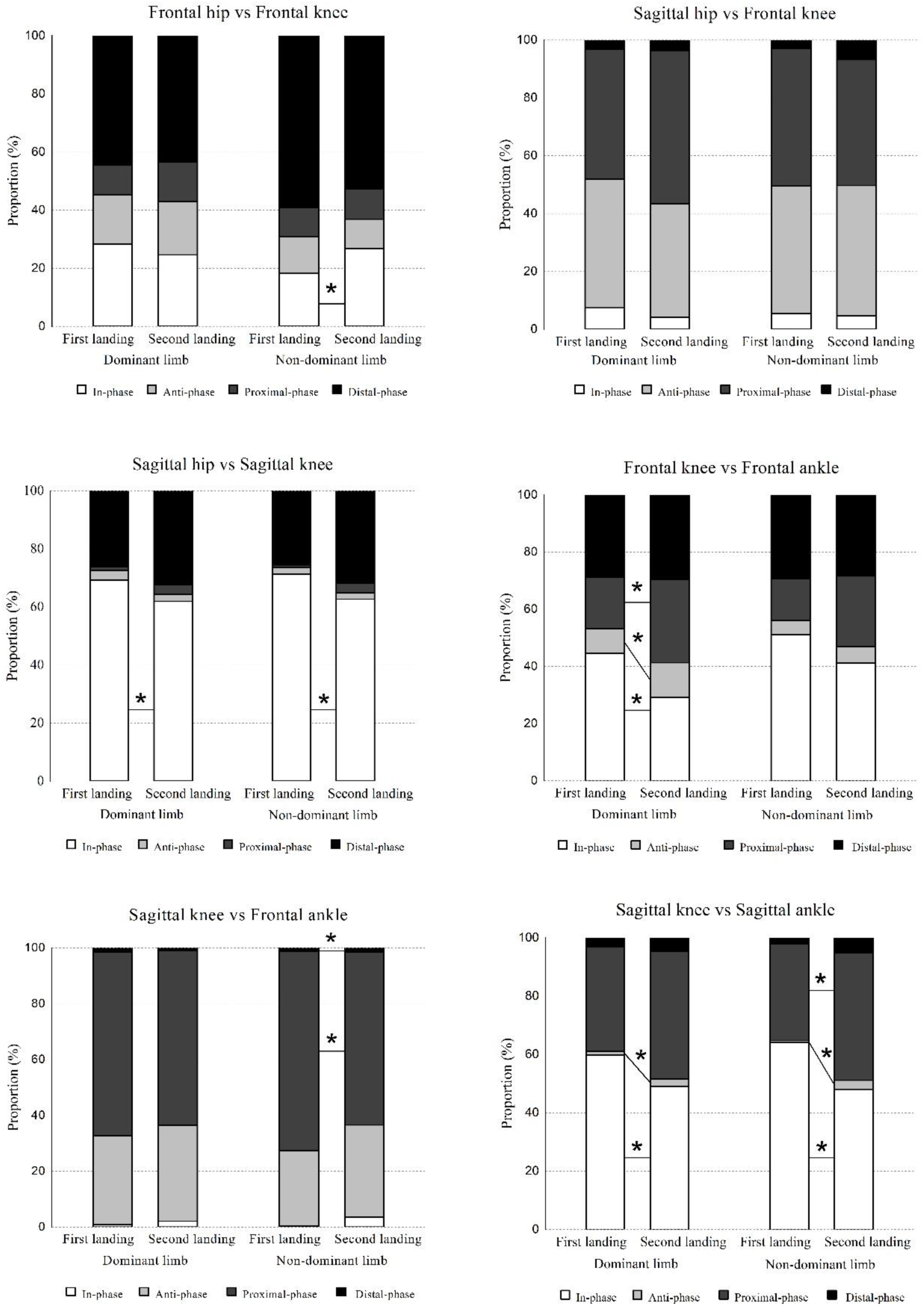
Coordination patterns of lower limb joints in different landing. Stacked graph for lower extremity joint coordination patterns during DJ task. The asterisk (*) indicates a significant change between the first landing and second landing (p < 0.05).

**Figure 2.**
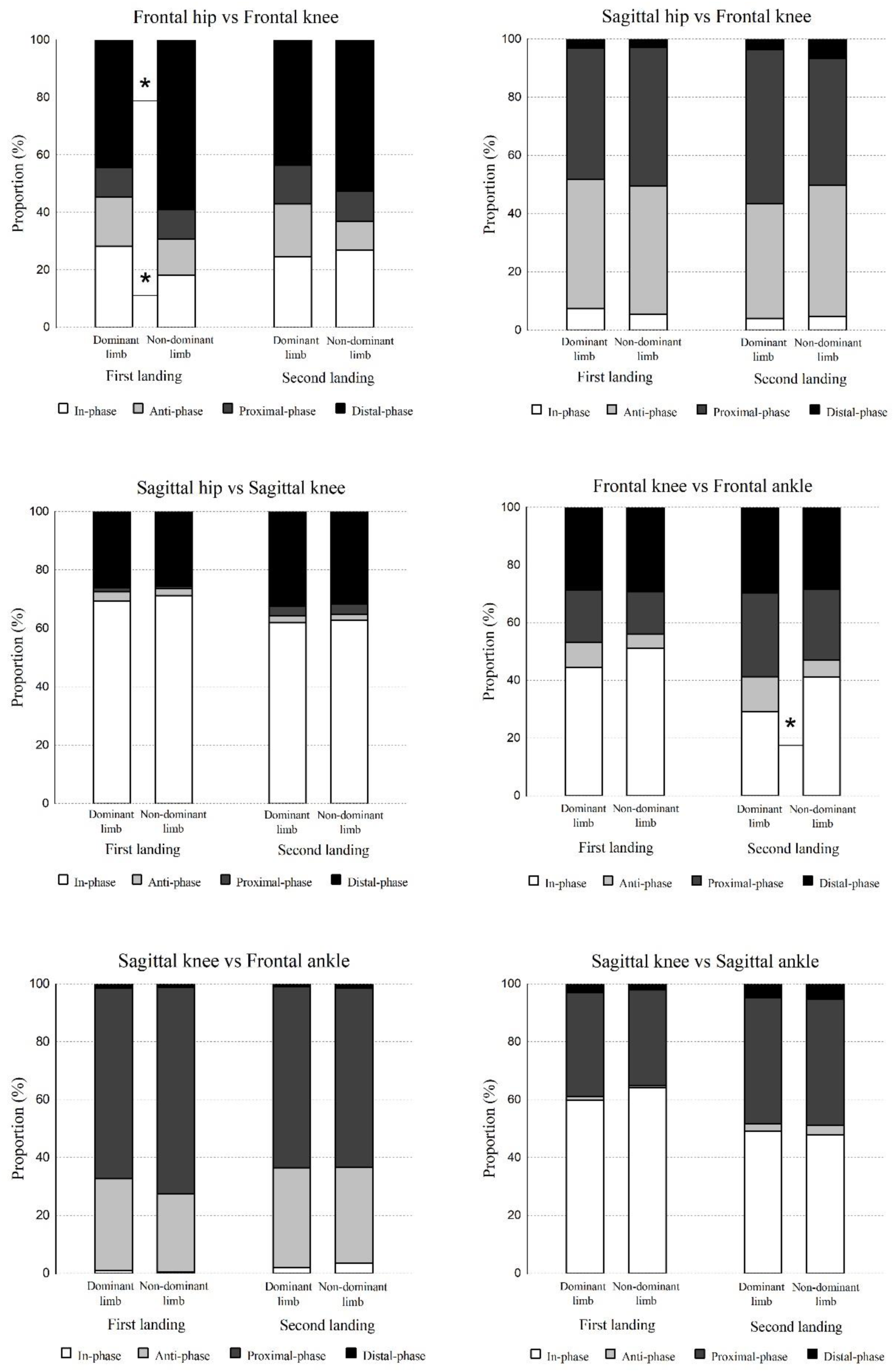
Side-to-side asymmetries of lower extremity joints in both landings of DJ task. The asterisk (*) indicates a significant change between the Dominant limb and Non-dominant limb (p < 0.05).

For HsKs couples, coordination patterns were concentrated in in-phase (where the hip and knee joints rotate in the same direction with similar amplitudes) on two landing phases. A significant main effect for the landing factor on in-phase (p < 0.05). Specifically, during the second landing task, both the dominant limb (P = 0.048, ES = 0.69) and the nondominant limb (P = 0.035, ES = 0.79) exhibited a significantly less proportion of in-phase compared to the first landing. However, there were no significant coordination pattern differences between limbs during the same landing phase (p > 0.05).

For HsKf couples, coordination patterns were concentrated in proximal coordination (hip rotates dominantly compared to the knee) on two landing phases. No significant interaction effect was observed, and there were no significant main effects of landing phase or limb dominance on joint coordination patterns. The proportions of the four coordination patterns did not show significant differences (P > 0.05).

For KfAf couples, coordination patterns were concentrated in in-phase (where the knee and ankle joints rotate in the same direction with similar amplitudes) on two landing phases. There were significant main effects for the landing factor found for the coordination patterns of anti-phase, in-phase and proximal phase (p < 0.05). Specifically, on the dominant limb, the proportion of in-phase during the first landing is significantly higher than second (p = 0.10, ES = 0.97). In contrast, the proportion of anti-phase (p = 0.013, ES = –1.2) and proximal phase (p = 0.018, ES = –1.07) are significantly lower during the first landing compared to the second landing. Additionally, there is a significant main effect of the limb dominance factor on in-phase (p < 0.05), it was found that on the dominant limb during the second landing, the proportion of in-phase is significantly less than on the nondominant limb (p = 0.013, ES = –0.73).

For KsAf couples, coordination patterns were concentrated in proximal phase (knee rotates dominantly compared to the ankle) on two landing phases. There were significant main effects of the landing factor on proximal and distal phase proportions (p < 0.05). Specifically, during the first landing task, the nondominant limb exhibited a significantly higher proportion of proximal phase compared to the second (P = 0.031, ES = 0.98). In contrast, the proportion of distal phase was significantly lower during the first landing phase compared to the second landing (P = 0.024, ES = – 0.91).

For KsAs couples, coordination patterns were concentrated in in-phase on two landing phases. There were significant main effects of the landing factor on anti-phase, in-phase, and proximal phase (p < 0.05). Specifically, the proportion of in-phase during the second landing was significantly lower compared to the first for both limb (P < 0.05), while the proportion of anti-phase was significantly higher during the second landing than the first (P < 0.05). Furthermore, on the nondominant limb, the proportion of proximal phase during the first landing was significantly less than the second landing (P = 0.024, ES = –0.68).

The variability of lower extremity joint coordination across different landing phases is illustrated in Figure 3. Importantly, for all HsKs, KsAs, KfAf, and KsAf couples, there were significant main effects of the landing factor on CAV (P < 0.05). Specifically, the variability of coordination during the first landing was significantly lower on both limbs compared to the second (P < 0.05). However, there were no significant main effects of limb dominance factor on any of the CAV measures (P > 0.05).

**Figure 3.**
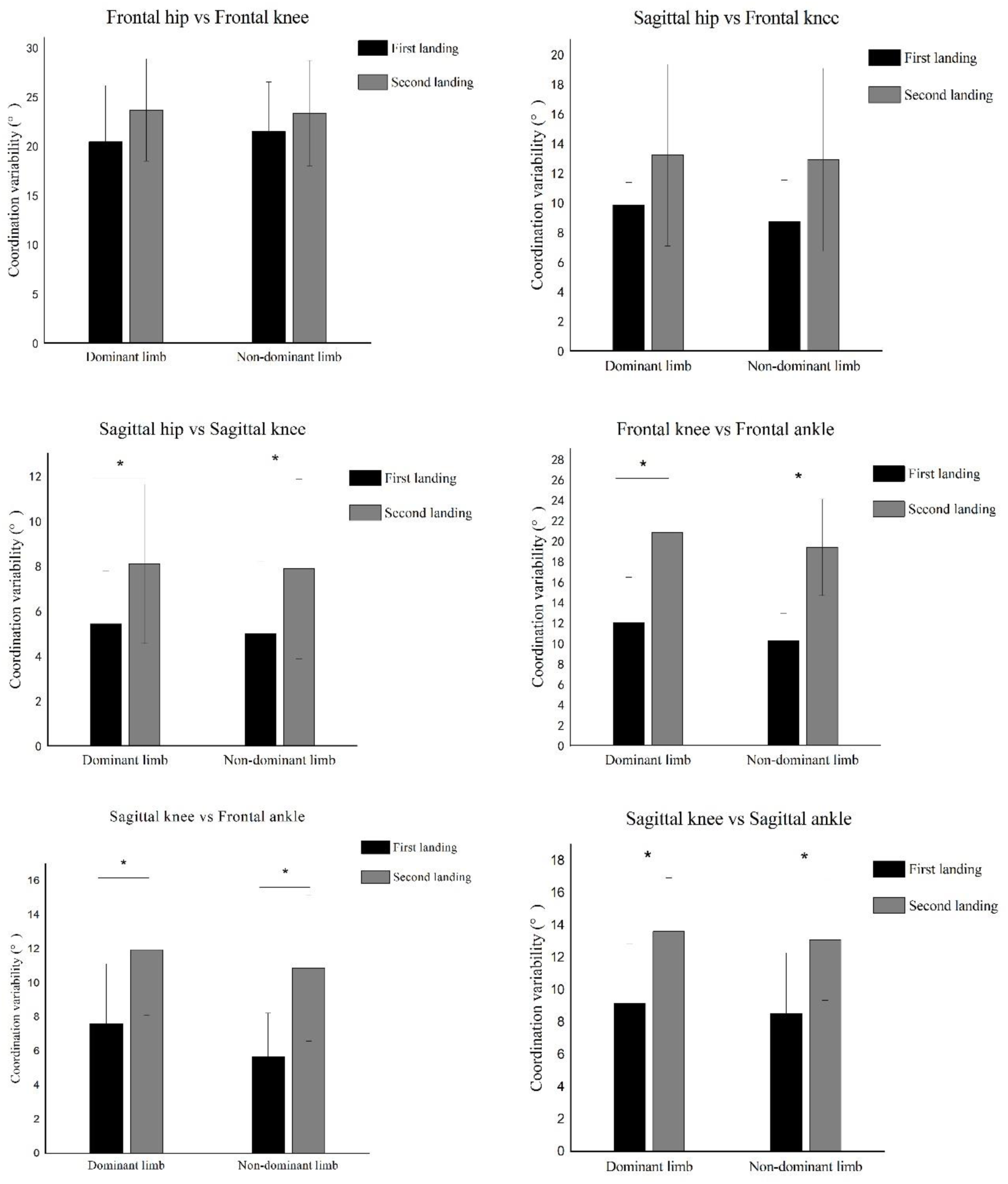
CAV of lower extremity joints at different landing. Stacked graph for lower extremity joint CAV during DJ task. The asterisk (*) indicates a significant change between the first landing and second landing (p < 0.05).

## 4 Discussion

The present study aimed to investigate whether there are differences in lower extremity joint coordination patterns and variability between the two landing phases during the DJ task. The results revealed that the proportions of each coordination pattern on both limbs changed in both landings. Specifically, the in-phase decreased, while the anti-phase increased during the second landing compare with first. Moreover, the coordination variability during the second landing was consistently higher than first. However, there were no significant differences in bilateral asymmetry between the two landing phases, which supported some of our research hypotheses. Previous research has indicated that changes in coordination patterns and increased coordination variability are related to injury during landing^10, 24^. This study is the first to investigate lower extremity joint coordination patterns and variability during the DJ task by comparing the two landing phases.

Through modified vector coding technique of hip and knee motions in the sagittal plane, it was found that compared to the first landing, both limbs showed a decrease in in-phase during the second landing, indicating reduced synchronization of hip and knee flexion motions during the second landing. A previous study reported that the second landing resulted in smaller flexion angles at the hip and knee^3^, suggesting that participants adopted a more upright posture during the second landing, leading to increased GRF on the lower extremity joints and an elevated risk of ACL injuries. Regarding the frontal-plane motions at the hip and knee, the second landing exhibited more in-phase, indicating enhanced consistency in hip and knee adduction/abduction motions. This may serve as a protective strategy since the reduction in sagittal-plane coordination increases the risk of injuries, leading to a tendency towards greater consistency in frontal-plane motions.

In this study, compared to the first landing, the KsAf couples demonstrated a decrease in proximal phase (knee dominance) and an increase in distal phase (ankle dominance) during the second landing. This suggests that in the second landing, the sagittal-plane knee motion decreased, while the frontal-plane ankle motion increased. DeLeo et al. suggested that altered joint coordination results are caused by variations in the relative timing or amplitude of motion^25^. Therefore, the observed changes in coordination in this study may also result from alterations in the alternating joint motion amplitudes, alternating joint motion timing, or a combination of both. Specifically, the decreased knee motion observed in KsAf enhances the consistency with the coordination in the frontal-plane hip motion, while also being associated with an increased demand for the frontal-plane ankle motion. This is analogous to the mechanism observed in individuals with ankle instability during jumping or landing tasks. In individuals with ankle instability, the increased frontal-plane motion during jumping tasks elevates the risk of joint instability during landing^26^.

An interesting observation from the present study was that during the coordination of knee and ankle joints in the sagittal and frontal planes, the second landing exhibited reduced in-phase, increased anti-phase, and a trend of increased proximal phase (knee dominance) compared to the first landing. The decreased proportion of in-phase in both sagittal and frontal plane motions indicates a mismatch in the flexion and inversion/eversion amplitudes between the knee and ankle joints, resulting in asynchronous of two joints during the second landing. Excessive motion of the lower extremities while landing contributes to overuse injuries, and anti-phase motion may overload the soft tissues or joints^21^. Therefore, alterations in the proportion of in-phase and anti-phase during the second landing may heighten the risk of knee joint and ankle joint injuries. The observed results of greater proximal (knee) dominancy during the second landing were likely due to increased knee motion while performing dynamic tasks compared with that in the first landing. As such, switching to second landing will lead to a faster rotating rate of knee flexion and inversion/eversion than those of the ankle, indicating that more load was transferred to the knee joint and less to the ankle joint. The heightened knee motion in the frontal plane during the second landing may elevate the risk of ACL injuries^27^. The differences in coordination patterns between the knee and ankle joints during the two landing may be attributed to the contrasting flight times preceding each landing. Specifically, the second landing benefits from a longer flight time due to the ascent and descent of the CoM, whereas the first landing only involves the descent of the CoM. This extended flight time preceding the second landing potentially allows for greater preactivation of the larger muscle groups, including the hamstrings^28^. This enables the body to better attenuate the energy when landing from a height. Additionally, some studies have reported that in adults, compared to the ankle joint, the knee joint absorbs greater amounts of energy with the moments at the knee joint when landing from a height^2^. Therefore, during the second landing, more anti-phase and knee dominant motion are observed. Overall, the increase knee flexion was surmised to compensate for deficits in ankle dorsiflexion and stabilize the ankle by increasing frontal plane motion, effectively absorbing kinetic energy during landing. However, excessive flexion and inversion/eversion motion may elevate the risk of knee injuries. Nevertheless, the exact underlying reasons for the observed phenomena in this study need further analysis, including research on muscle activation patterns and morphological changes during the two landing phases in the DJ task.

The observed side-to-side asymmetries during the dynamic task have been suggested as a precursor to non-contact ACL injuries^29^. In this study, we only observed significant asymmetry in the HfKf and KfAf couples. Moreover, during the two landing phases of the DJ task, most couplings showed no significant differences in joint coordination asymmetry. Unlike previous findings, Bates et al. found great bilateral asymmetry in the hip and knee kinematics and kinetics for both the frontal and sagittal planes during the second landing compared to the first. For instance, the second landing exhibited increased side-to-side asymmetry for hip sagittal and horizontal plane rotation angles and knee flexion angles^3^. Potential reasons for these differences could be attributed to two factors: firstly, the differences in the gender of the participants, as female athletes are considered to exhibit greater side-to-side asymmetry during bilateral landing tasks compared to males^29^; secondly, the variations in the analysis techniques, as this study employed non-linear analysis techniques, which are more sensitive in detecting subtle variations in kinematic data compared to linear analysis^30^.

The variability of coordination is often considered to be related to the neuromuscular control function of the human body^16, 31^. In this study, several joint couples, especially between the knee and ankle joints in multiple planes, showed significant differences in coordination variability between the first and second landing phases. Compared to the first landing, the second landing exhibited greater coordination variability, indicating the adoption of different motor strategies between the two landings. Dynamic systems theory describes the range of variability as a “bell curve”^32^, where excessively low variability may be associated with task maladaptation, while high variability may suggest insufficient coordination or sensory-motor control capabilities^31^. However, in some studies, high variability in healthy individuals is considered an indication of better adaptability to several situations, reflecting a higher level of flexibility in the neuromuscular system^33^. Therefore, healthy window of variability has not yet been defined. In dynamic tasks like the DJ, it is difficult to determine the optimal range of coordination variability for both flexibility and control. However, considering the stability of the testing environment, the consistency of the testing task, and the significant increase in anti-phase proportion in multiple joint couplings during the second landing compared to the first landing, the increased variability observed during the second landing in the DJ task likely reflects weaker lower extremity motor control rather than the flexibility of the neuromuscular system. Previous studies on coordination variability in various jumping tasks have indicated that higher variability in healthy individuals is associated with an increased risk of sports injuries^18, 19^. From this point of view, the increased coordination variability may represent aberrant neuromuscular motor control.

Understanding the mechanisms underlying the variability changes during the two landing phases of DJ task is a crucial focus for future research. Previous studies have suggested that differences in external focus could be one of the factors contributing to the coordination changes^24^. During the first landing, participants receive instructions about the landing spot and sound, directing their attention towards leg mechanics, which helps reduce the impact force and lower body stiffness during landing^34^. However, between the vertical jump and the second landing, participants do not receive any external cues, leading to a direct alteration in muscle activity patterns. In the first landing, muscles are activated not only to counteract the impact GRF but also to generate power for a vertical jump, while in the second landing, muscle activation is primarily aimed at countering the GRF^9^. Consequently, compared to the first landing, the body experiences increased perturbations before the second landing, making posture stability more challenging to control.

This study had several limitations. First, the research only measured the coordination of adult males during landing in DJ task, whereas female athletes have reported 4-6 times higher incidence of ACL injuries compared to males^35^. Therefore, to develop a more comprehensive understanding of the relationship between ACL injuries and coordination during landing, future studies should consider including female participants. Second, while all participants had jumping and landing experience, variations in their skill and experience levels may have influenced the biomechanical characteristics during landing. Therefore, in future investigations, it would b e beneficial to consider the participants’ skill and experience levels in more detail to understand how these factors influence coordination changes.

## 5 Conclusion

Our research findings indicate that during the execution of the DJ task, there are changes in joint coordination patterns and an increase in coordination variability in the second landing compared to the first landing, particularly evident in the couples of the knee and ankle joints. This reflects a reduction in lower extremity neuromuscular control during the second landing. Therefore, as observed in this study, the second landing may pose an increased risk of injury to the knee and ankle compared to the first landing. These findings are significant for understanding the mechanisms of sports injuries and implementing appropriate preventive measures.

### Practical Applications

The assessment of the two landing phases in the DJ task reveals crucial information about the joint coordination patterns adopted by male athletes. In the second landing, the knee and ankle exhibit less favorable coordination strategies, with increased variability that raises the risk of joint injuries. These findings contribute to a better understanding of the mechanisms underlying injury occurrence and provide insights into effective preventive measures for athletes. Specifically, it is recommended to implement resistance training methods, including plyometric exercises, to enhance muscle strength. Additionally, during the second landing, coaches can employ visual and auditory cues to direct athletes’ attention towards the knee and ankle, promote soft landings, increase knee flexion angles upon landing, and incorporate strategies such as jumping to another box to reduce impact forces during the second landing of the DJ task. These interventions aim to improve potentially harmful mechanical patterns.

## Data Availability

Data available on request from the authors: The data that support the findings of this study are available from the corresponding author upon reasonable request.

## Acknowledgment

The authors would like to express their gratitude to all study participants.

